# Accessioning and automation compatible anterior nares swab design

**DOI:** 10.1101/2020.09.24.20200386

**Authors:** Mary E. Pettit, Sarah A. Boswell, Jason Qian, Richard Novak, Michael Springer

**Author notes:** Corresponding author. (M.S.). These authors contributed equally to this work (Mary E. Pettit, Sarah A. Boswell).

## Abstract

The COVID-19 pandemic has resulted in an unparalleled need for viral testing capacity across the world and is a critical requirement for successful re-opening of economies. The logistical barriers to near-universal testing are considerable. We have designed an injection molded polypropylene anterior nares swab, the RHINOstick, with a screw cap integrated into the swab handle that is compatible with fully automated sample accessioning and processing. The ability to collect and release both human and viral material is comparable to that of several commonly used swabs on the market. SARS-CoV-2 is stable on dry RHINOstick swabs for at least 3 days, even at 42°C, and elution can be achieved with small volumes. The swab and barcoded tube set can be produced, sterilized, and packaged at < 2 USD per unit and can easily be adopted by large research institutes to increase throughput and dramatically reduce the cost of a standard SARS-CoV-2 detection pipeline.

## INTRODUCTION

As of September 8^th^ 2020, at least 27 million cases of COVID-19 and 890,000 deaths have been reported world-wide (1). To determine if a patient has COVID-19, in most cases, a nasopharyngeal (NP) swab is collected by a trained professional. The swab is then deposited in 1-3 mL of transport media followed by RNA purification and RT-qPCR. NP swabs are around 15 cm in length with a collection head coated with short synthetic filaments, flock, or spun fibers (2); collection is often an uncomfortable process. The high demand for testing during this pandemic has outstripped the supply of NP swabs (and many other critical reagents for testing) resulting in a testing bottleneck (3). These supply limitations together with a drive towards patient self-collection has spurred the development of alternatives to the standard NP swab. A promising alternative is anterior nares (AN) swabs, commonly referred to as nasal swabs. AN swabs offer a testing sensitivity similar to NP swabs (4, 5) but are easier to use and more comfortable for the patient.

The choice of swab and collection device can have a major impact on the testing speed in clinical labs. Upon receiving samples, a typical procedure in a testing facility is to first accession the delivered patient samples by scanning the barcoded label to upload relevant patient data into the system, then swabs are manually removed from each collection tube and disposed of. The sample transport media is then processed to purify RNA, which is used as input for RT-qPCR. The initial steps in this procedure are hard to automate, slow, and expose staff to infection. Standard 1D barcoding systems and the manual removal of swabs is time consuming and thus costly. There are machines that can perform the entire procedure from accessioning to results, one tube at a time, e.g. a cobas® 8800, but this process is slow, 1056 tubes per 8-hour shift (6), and the machines are expensive.

In an effort to meet the dramatic increase in demand for nasal swabs, several groups have designed and 3D printed new swabs (2, 7). The performance of these swabs is comparable to that of standard swabs; however, they aim to reproduce the existing status quo, rather than to address some of the limitations caused by the standard swab design. An ideal swab would be one that is comfortable for patients to self-administer without sacrificing performance, while also allowing for automated specimen accessioning and processing. Additionally, the swab would be made from non-absorbent material, allowing samples to be diluted into smaller volumes of transport media than those used in the current procedure, rendering the sample more concentrated and allowing for more sensitive detection of viral RNA. Here we present the RHINOstick, a swab that 1) performs as well as existing AN swabs; 2) is compatible with direct input to RT-qPCR for extraction free SARS-CoV-2 detection; and 3) is compatible with a collection system (swab and tube) that enables automated sample accessioning and processing.

## MATERIALS AND METHODS

### Swab design

The swabs were designed in SolidWorks (Dassault Systèmes) and manufactured using single-shot rapid injection molding (Protolabs) from medical grade FHR P5M4R polypropylene (Flint Hills), a material compatible with autoclaving (121°C, 20 min), ethylene oxide, gamma radiation, and e-beam sterilization. The stacked rings of the swab head enable collection of nasal matrix without the need for an absorbent coating, using a design previously developed at the Wyss Institute. The cap cavity is compatible with automated decapping robot systems using a square profile adapter head, while the 2 mm pitch external threading mates with the interior threading of sample collection tubes from several major manufacturers (e.g. Matrix, Micronics, and LVL). As the new swab we developed in this study is useful for the collection of nasal samples for diagnostic tests we call it the RHINOstick swab.

### Absorption of liquid by swab

The swabs used in this study were weighed on an analytical balance before and after a 15 second incubation in 1 mL of nuclease free water. Six replicates were measured and results are reported in Table 1.

**Table 1:**

| Swab | Collection Material | Average volume absorbed (µL) | Standard deviation (µL) | Purchasable as of 09/17/20 |
| --- | --- | --- | --- | --- |
| **RHINostick** | Polypropylene | 14.4 | 2.2 | N |
| **P&G blue** | Polypropylene | 0.7 | 1.0 | N |
| **Wyss flock** | Polypropylene and polyester flock | 65.8 | 3.9 | N |
| **Puritan hydraflock** | Polyester flock | 154.1 | 8.9 | Y |
| **Puritan foam** | Polyurethane foam | 41.3 | 14.4 | Y |
| **Puritan polyester** | Polyester | 155.9 | 9.6 | Y |
| **US Cotton** | Cotton | 168.8 | 25.4 | Y |
| **Microbrush** | Nylon Flock | 64.9 | 10.1 | Y |

### Anterior nares self-swabbing to compare swab performance

We compared several swab types for performance in anterior nares (AN) specimen collection: the RHINOstick prototype, Proctor & Gamble (P&G) blue prototype, a Wyss Institute flocked prototype, Puritan hydraflock, Puritan foam,

Puritan polyester, US Cotton, and Microbrush®. Per CDC guidelines, volunteers were instructed to insert the swab 0.5 inch into a nostril, rotate three times along the membrane of the nose firmly and leave in place for 10 to 15 seconds, remove, and then repeat this procedure on the other nostril with the same swab to collect nasal matrix (8). The volunteer was then instructed to place the used swab in a dry 1.5 mL microcentrifuge tube and break the handle if necessary so the tube could close for transport. Prior to RT-qPCR reactions all swabs were suspended in 200 uL of nuclease free 1x PBS. All experiments in this study were approved by the Institutional Review Board of the Harvard Faculty of Medicine, IRB20-0581, and informed written consent was obtained from volunteers.

### RT-qPCR

RT-qPCR reactions were prepared to reach a final volume of 10 µL using 8 µL of master mix and 2 µL of sample. The Luna Universal One Step RT-qPCR kit (NEB) was used for all RT-qPCR reactions. The master mix protocol was adjusted to include 0.25 U/µL of RNaseIn Plus (Promega) for every 10 µL reaction. RT-qPCR reactions were run on the QuantStudio 6 Real Time PCR system (Thermo Fisher Scientific) following the manufacturer recommended Luna RT-qPCR protocol. For all reactions, melt curves were used to determine if products were specific or non-specific. All non-specific Tms, >0.5 °C from the expected melting temperature, are presented as having a Ct of 40. All experiments included at least one negative control which was either 1x PBS or water. The sequences of all primers used are listed in Table S1.

### Recovery of human mRNA from AN swabs

SARS-CoV-2 negative volunteers performed AN swabbing as directed with each type of swab tested (Fig. 2, Table 1) to collect nasal matrix. There were three biological replicates for each AN swab measurement, taken on at least two different days. For every condition in which a swab was tested, an unused swab, without nasal matrix, was processed in parallel as a negative control. To recover the sample from the swabs, all swabs were suspended in 200 µL of 1x PBS, vortexed for 10 sec, spun down in a microcentrifuge, and input directly to the RT-qPCR for GAPDH mRNA detection (Fig. 2C).

**FIG 1.**
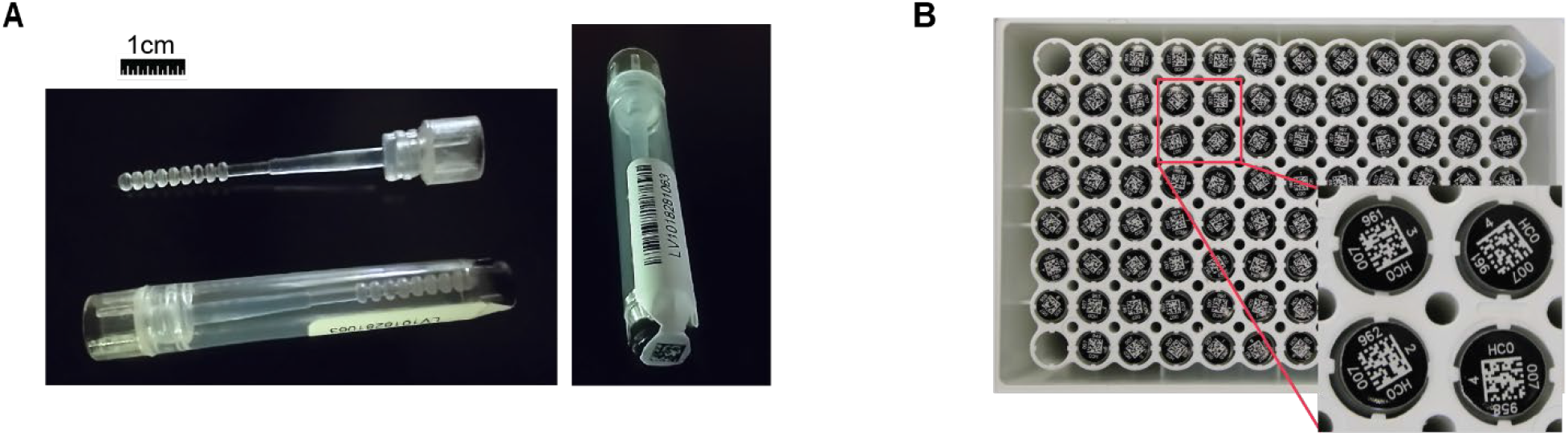
96-well format automation and accession compatible AN swab design. (A) Custom injection molded AN swab that can be produced at large scale and is compatible with 96-well format automation. A sample tube compatible with the RHINOstick swab is shown with barcodes on the side and bottom. The RHINOstick swab is 4.9 cm long with a collection head length of 1.6 cm. 1 cm scale bar shown for reference. (B) 96-well rack of swabs and tubes with 2D matrix codes printed on the bottom of the tubes, allows for rapid accessioning.

**FIG 2.**
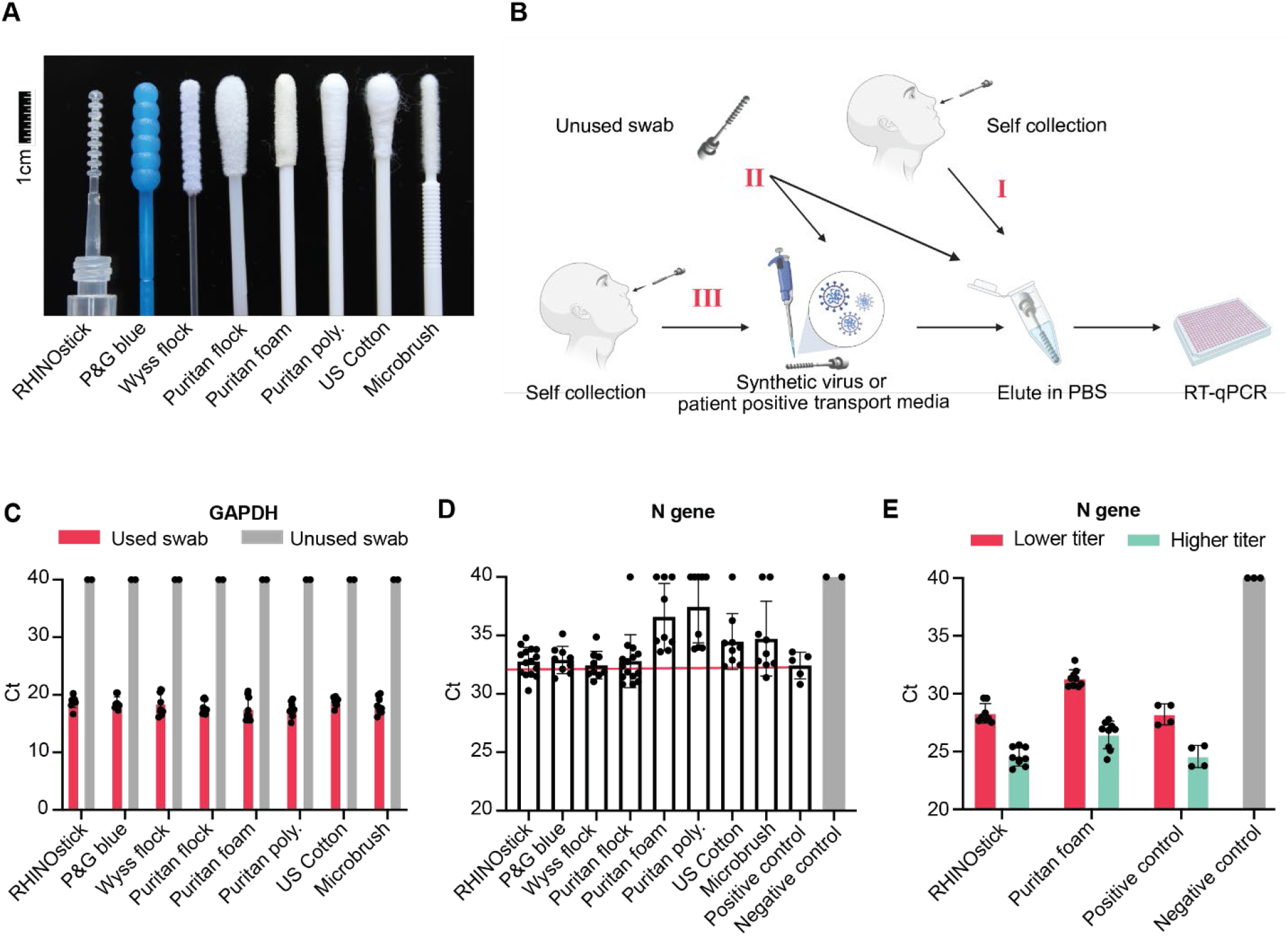
Comparison of swab performance. (A) AN swabs tested in this study, from left to right: RHINOstick, Proctor & Gamble (P&G) blue, Wyss Institute flocked prototype, Puritan hydraflock, Puritan foam, Puritan polyester, US Cotton, and Microbrush®. 1 cm scale bar shown for reference. (B) Schematic of swab experiments performed in C-D. **I**; SARS-CoV-2 negative volunteer self-collected nasal matrix on a swab. **II**; unused swab, without nasal matrix, was either treated with packaged synthetic SARS-CoV-2 virus or left untreated (clean, unused swab). **III**; SARS-CoV-2 negative volunteer self-collected nasal matrix on a swab which was then treated with packaged synthetic SARS-CoV-2 or SARS-CoV-2 clinical sample (Methods). All samples were eluted in PBS and used as direct input to RT-qPCR assays. Images created with BioRender.com. (C) RT-qPCR quantitation of human GAPDH mRNA from used swabs containing nasal matrix (pink bars) or matched unused swabs (grey bars). (D) RT-qPCR quantitation of the SARS-CoV-2 N gene from packaged synthetic virus applied to clean, unused swabs. The grey bar is the negative control, PBS input into RT-qPCR. The pink line is a guideline for complete recovery based on the positive control. (E) RT-qPCR quantitation of SARS-CoV-2 N gene from swabs in the presence of nasal matrix spiked with a lower (∼140 copies/µL, pink bars) or higher (∼1600 copies/µL, green bars) titer clinical sample. The grey bar is the negative control, PBS, and the positive controls are the lower or higher titer clinical samples directly input to RT-qPCR. RT-qPCR data in C-E all show technical replicates of at least 3 biological experiments.

### Contrived samples using packaged synthetic SARS-CoV-2 spiked onto unused swabs

AccuPlex SARS-CoV-2 verification panel v2 (Seracare), a packaged synthetic virus, containing the N gene, E gene, ORF1a, S gene, and RdRp was used to simulate the expected viral recovery from AN swabs near the limit of detection (Fig. 2D, S2C). 10 µL of 100 copies/µL packaged synthetic virus was directly applied to the collection head of each swab. Swabs were left in a fume hood for about 20 min until the swabs appeared dry to the eye indicating absorption of the packaged synthetic virus into the collection material. At least three biological replicates were used for every swab tested and replicate data was collected on at least two different days. Swabs were then inserted into a 1.5 mL microcentrifuge tube containing 200 µL of 1x PBS, vortexed for 10 sec, spun down in a microcentrifuge, and 2 µL was input directly to RT-qPCR for N gene detection. The positive control was 10 µL of 100 copies/µL packaged synthetic virus directly input to 190 uL of PBS.

Clinical samples

NP swabs from SARS-CoV-2 patient samples were purchased from BocaBiolistics, FL. The NP swabs are remnant samples obtained through BocaBiolistics and partner labs that were de-identified by BiocaBiolistics with their IRB reviewed and approved SOP for de-linking specimens. These NP swabs arrived in 1-3 mL of viral, multitrans, or universal transport media (VTM, MTM, or UTM). 40 µL of each sample was aliquoted and frozen at -80°C to limit freeze-thawing of samples.

### Contrived samples from a clinical source spiked onto swabs with nasal matrix

Nasal matrix was collected from volunteers as described above using RHINOstick and Puritan foam swabs. 5 µL of clinical sample, with either a higher (∼1600 copies/µL), or lower titer (∼140 copies/µL), were applied to the collection head of used swabs, and swabs were air dried in the BSL2+ biosafety cabinet for 20 min. Each swab was then placed in a 1.5 mL microcentrifuge tube containing 200 µL of 1x PBS, manually spun for 10 sec in the media, and 2 µL was directly input to the RT-qPCR for both N gene (Fig. 2E) and GAPDH mRNA detection (Fig. S2C). To assess maximum possible viral recovery from the swab, the positive control was 5 µL of either the higher or lower titer clinical sample in 195 µL of 1x PBS. Negative controls are unused RHINOstick and Puritan foam swabs suspended in 200 µL of 1x PBS. Three biological replicates were performed for each titer and type of swab tested.

### Assessment of stability of SARS-CoV-2 on swabs with nasal matrix over time

To assess the stability of the SARS-CoV-2 virus on swabs with nasal matrix over time, two volunteers self-swabbed three independent times with both the RHINOstick and Puritan foam swabs for a total of six swabs at each time point. The handles of the Puritan foam swabs were broken in order to safely close the collection vial, a 1.5 mL microcentrifuge tube. Several clinical samples were mixed together to generate a pooled clinical sample with a viral titer of ∼10,200 copies/µL. The pooled clinical sample was then aliquoted into 50 µL volumes and refrozen at -80°C. At each time point (72, 48, 24, 2, and 0 hrs) an aliquot was thawed and 3 µL of pooled clinical sample was applied to each swab. One RHINOstick and one Puritan foam swab with nasal matrix from each volunteer was incubated dry at room temperature (25°C) or 42°C in a 1.5 mL microcentrifuge tube to asses stability at room temperature or elevated temperatures that may occur during transport. A matched RHINOstick or Puritan foam swab with nasal matrix from each volunteer was immediately put into a 1.5 mL microcentrifuge tube containing 0.4 mL of 1x PBS to assess the relative stability of a wet swab vs dry swab. An additional 3 µL of the pooled clinical sample was applied to an unused RHINOstick and unused Puritan foam swab at each time point and kept dry over the time course at 25°C, to assess the effect of nasal matrix on viral recovery. At the end of the time course, dry swabs were suspended in 0.4 mL of 1x PBS. The samples from both wet and dry tubes were mixed by vortexing for 10 sec, then spun down in a microcentrifuge. 2 µL of each sample was directly input to RT-qPCR for GAPDH and N gene detection. The positive control was 3 µL of the pooled clinical sample in 197 µL of 1x PBS at time 0.

## RESULTS

### Swab design for automated accessioning and analysis

NP swabs are long, making it challenging to use these swabs with automation-compatible tubes. AN swabs in contrast do not need to be as long as NP swabs and can be designed with a shorter handle, opening up the possibility of making AN swabs that can be directly paired with automation-compatible tubes for an effective collection system. As part of the design, RHINOstick swabs have a cap that can be directly screwed onto a 96-well format automation-compatible tube, such as a 1.0 mL Matrix tube (Thermo Fisher Scientific) (Fig. 1A). The swabs were made by single shot injection molding with medical grade polypropylene (Fig. S1 and Methods). Injection molding of swabs allows for high volume production at low prices. While the swabs can fit onto many tubes, we believe the optimal design is in collection tubes pre-labeled (by the manufacturer) with a serialized Type 128 1D barcode plus human readable code on the side with a matching 2D data matrix barcode on the bottom. (Fig. 1). This design allows for the collection tube and swab to be accessioned and used by the patient in an unobserved manner without having to pre-register each barcode manually, reducing costs and labor. In addition, the matching 2D barcode on the bottom allows a whole rack of tubes to be accessioned in seconds by a barcode reader.

### Swab performance

We compared the RHINOstick to several other swabs on the market or under development (Fig. 2A, Table 1). First, we tested for absorption of water. Water absorption is sometimes used as a proxy for the amount of material that a swab will collect (9, 10), although it does not necessarily correlate with effective collection of cells and viral particles. The RHINOstick, as well as the Proctor and Gamble (P&G) blue swab absorbed very little water compared to the majority of available swabs on the market or prototypes. This lack of absorption is likely because polypropylene is more hydrophobic than the other collection materials, such as cotton and spun polyester.

To test swab performance more directly, we measured the performance of 8 different AN swabs using several approaches (Fig. 2). We tested collection and recovery of 1) human mRNA in nasal matrix from swabs, 2) mRNA from viral particles added to swabs, and 3) mRNA from viral particles added to swabs coated in nasal matrix (Fig. 2B). Human mRNA was used as a process control to assess successful collection and recovery of cells from swabs. The process control also assesses the efficiency of the reverse transcription (RT) reaction as the primers span two exons to ensure the assay quantifies mRNA not DNA (11). A single volunteer swabbed with 8 different brands of AN swabs in triplicate (Fig. 2B, scheme I) and the eluent was used as direct input for RT-qPCR for GAPDH mRNA to quantify the amount of human mRNA recovered (Fig. 2C). All 8 swabs performed similarly in this assay and no GAPDH was detected on any of the unused swabs (Fig. 2C). For all evaluations of AN swabs in this work we performed direct RT-qPCR on the swab eluant without RNA purification.

Recovery of viral particles was first assessed on a contrived sample by applying packaged synthetic SARS-CoV-2 viral particles (Seracare reference) to an unused swab for each of the 8 AN swabs tested (Fig. 2B, scheme II). The packaged synthetic virus was dried onto the swab and eluted into PBS by vortexing. In a similar experiment, we found that elution into PBS by gentle swirling of the swabs releases the virus at equivalent or superior levels to vortexing in the same amount of time (Fig. S2A and B). The level of viral particles released by each swab was quantified by RT-qPCR for the SARS-CoV-2 N gene (Fig. 2D). The RHINOstick performed as well as the other swabs tested, and released an equivalent number of viral particles to the positive control (Fig. 2D). The lower detection of viral RNA for other swabs such as the Puritan foam is likely due to the fact that these swabs absorb significant volumes of liquid (Table 1) making it hard to elute the contents off the swab efficiently, especially given that the maximal recovery of AccuPlex possible is 10 molecules per reaction. Going forward, due to swab availability, and its common use we performed all comparisons to the RHINOstick with only the Puritan foam swabs.

To test recovery of SARS-CoV-2 RNA from contrived clinical samples in the presence of nasal matrix, volunteers self-swabbed using either the RHINOstick or Puritan foam swab, then transport media from SARS-CoV-2 clinical samples was applied to the used swabs (Fig.2B, scheme III). After drying, the viral material was recovered by spinning the swabs in PBS. This experiment was performed with both a lower and a higher titer clinical sample (Methods), and the presence of both SARS-CoV-2 N gene and GAPDH mRNA was detected by RT-qPCR using the PBS/swab solution as direct input to RT-qPCR (Fig. 2E, Fig. S2C). Additionally, the equivalent performance of the RHINOstick to the positive control demonstrates the robustness of RT-qPCR to nasal matrix. The clinical sample titers were determined using an N gene standard curve (Fig. S2D, Supplementary Methods). RHINOstick swabs were not statistically distinguishable from the positive control at either titer, but the Puritan foam swabs showed lower recovery (P < 0.001 by an independent t-test). Replicate Ct values shows the high reproducibility of the qPCR data (Fig. S2E and F).

### Virus stability on swabs

A key issue with swabs is the stability of viral particles on the swabs during transport from the collection site to the test lab. To test the stability of SARS-CoV-2 on swabs over time we added SARS-CoV-2 from clinical samples to swabs containing nasal matrix (Fig. 3A). The contrived samples were left wet or dry at 25°C as well as dry at 42°C, to simulate storage in a hot car or truck, for up to 72 hours before elution into PBS. The presence of both SARS-CoV-2 N gene RNA and GAPDH mRNA was detected by using the swab eluent as direct input into RT-qPCR (Fig. 3 and S3).

**FIG 3.**
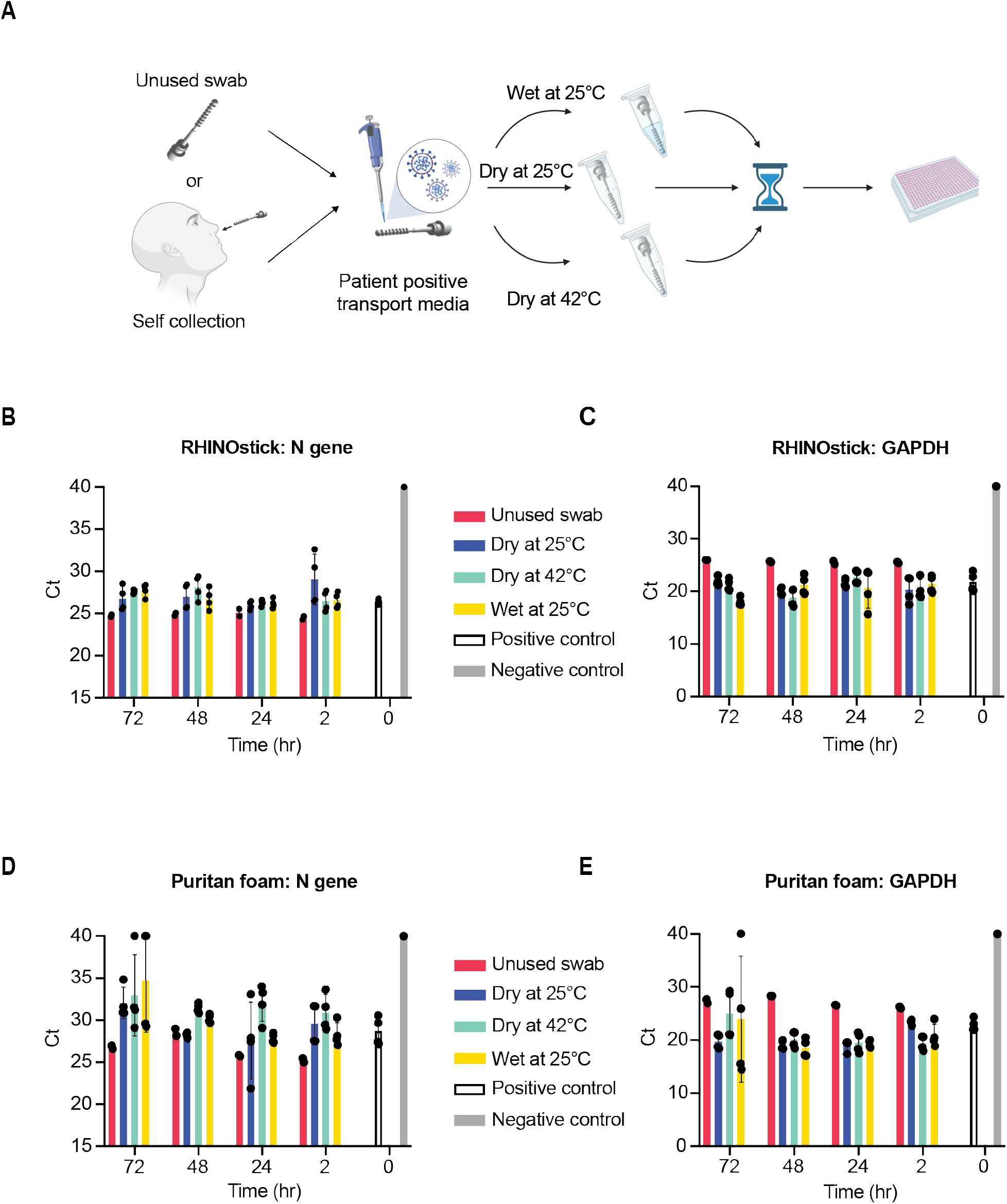
Stability of SARS-CoV-2 on swabs in the presence of nasal matrix. (A) Schematic of the experimental workflow in B-E. SARS-CoV-2 clinical sample was applied to unused swabs or self-collected AN swabs, with nasal matrix, (Methods) and left dry or wet at 25°C, for up to 72 hours. All samples were quantified by direct input of eluent into RT-qPCR. Images created with BioRender.com. (B,C) The stability of SARS-CoV-2 on RHINOstick swabs with nasal matrix left dry or wet at 25°C or dry at 42°C was analyzed over the course of 72 hours by RT-qPCR for the SARS-CoV-2 N gene (B) or GAPDH (C). (D,E) The stability of SARS-CoV-2 on Puritan foam swabs with nasal matrix left dry or wet at 25°C or dry at 42°C was analyzed over the course of 72 hours by RT-qPCR for the SARS-CoV-2 N gene (D) or GAPDH (E). Data points in B-E are technical replicates of 2 biological replicates. The positive control in B-E is the SARS-CoV-2 clinical sample directly added to PBS at time 0. The negative control is an unused RHINOstick (B, C) or Puritan foam (D, E) swab in PBS.

SARS-CoV-2 viral particles on the RHINOstick swabs were stable under all conditions tested both in the presence and absence of nasal matrix (Fig. 3B and S3A) whereas the Puritan foam swabs showed much greater variation in N gene detection when in the presence of nasal matrix, particularly when the sample was left out for 72 hours (Fig. 3D and S3A). Overall, GAPDH detection was more consistent for both the RHINOstick and Puritan foam swabs (Fig. 3C and D, S3B-D) across all conditions in the time course, but was slightly more variable for the Puritan foam swabs stored wet at room temperature for 72 hours (Fig 3E). The variability in the N gene as well as GAPDH data collected from Puritan foam swabs during the time course is also observed when comparing the Ct’s between two technical replicates in the RT-qPCR data (Fig. S3E and F).

## DISCUSSION

Our improved AN swab is comfortable to use, allows patients to perform swabs for themselves, and enables rapid accessioning and processing. The RHINOstick performs comparably to currently available swabs, releasing similar amounts of human and viral material into solution after use (Fig. 2). We found that RHINOstick and Puritan foam swabs detected similar levels of GAPDH mRNA (Fig. 3), while SARS-CoV-2 was detected more consistently from the RHINOstick swab with lower titer contrived samples (Fig. 2D and E) or after long periods of storage (Fig. 3). All RT-qPCR reactions performed in this study used direct input of swab eluant to the reaction mix without any RNA extraction and we were able to detect as low as 10 molecules per assay (Fig. 2D).

SARS-CoV-2 viral particles on the RHINOstick swab proved to be very stable with no statistically significant loss of Ct under all the conditions tested (Fig. 3). One of the key design elements of the RHINOstick swab is the ability for a patient to self-collect their AN swab for sample processing. To best use this feature, it is preferred to use dry swabs in which the swab is put into the collection tube after self-collection in the absence of any buffer. This swab may then be mailed in or collected at a central location without the need for concern over sample leakage in transport. The stability of SARS-CoV-2 on the RHINOstick swab for up to 72 hours before processing (Fig. 3B and C) demonstrates the feasibility of the dry swab method which is consistent with other studies (12). An additional advantage of the new swab design is the ability to elute the sample in a low volume of liquid (200 µL), potentially increasing the sensitivity of the direct RT-qPCR method by 5-15 fold compared to standard methods. Most commercial swabs cannot be used with this low elution volume, due to the high volume of liquid absorbed by the swab (Table 1).

We envision the RHINOstick swabs being used in the following workflow: the patient will scan the side of the barcode on the side of the tube using a cellphone app, phone-accessed website, or scanner and an ID card at the collection site to link the patient and sample together. After swabbing with the RHINOstick swab, the patient would screw the swab into the barcoded tube. The sample would then be packaged for Category B compliant transport. In an unsupervised self-collection setting, the tube could be rescanned at the sample deposition site to help track sample custody. The tubes would be deposited in a lockbox at the site, which would periodically be sent to the testing center. All swabs would be stored and transported dry avoiding the risk of liquid leakage. In the testing facility, the samples would be received and loaded into 96-well racks by hand (Fig. 1B). Each rack of tubes would then be put onto a robot that scans the 2D matrix codes on the bottom of the tubes thereby linking the sample ID to each plate and plate location in seconds. After accessioning, the samples can pass to a de-capping robot which removes the caps and the samples can then be eluted, inactivated, and processed for viral quantitation.

Here we demonstrate that the RHINOstick, a newly designed injection molded polypropylene swab with a screw cap integrated into the swab handle, performs equal to several commonly used AN swabs on the market at capturing and releasing SARS-CoV-2 viral particles from AN swabs. This innovative AN swab design has the potential to expedite SARS-CoV-2 diagnostic testing while significantly reducing costs. We anticipate that these swabs will be generally useful for pathogen panel testing at large research institutes.

## Data Availability

All data will be included in the supplementary material of the manuscript.

## ACKNOWLEDGEMENTS

We would like to thank Sam Keough, Raquel Arias-Camison, and Sandro Santagata for IRB and COMS help; Cepko lab for Twist Biosciences SARS-CoV-2 RNA; Chris Chidley for the design of the GAPDH primers used in this study; Rebecca Ward for review of this manuscript; and the volunteers who swabbed for this study. Funding: This work was supported by DARPA D18AC00006. JQ is supported by NSF GRFP.

## Author contributions

RN and MS conceived the study. MP and SAB performed most key experiments and data analysis (supervised by MS). JQ assisted with several experiments. SAB, MP, and MS wrote the paper. All authors reviewed the manuscript.

